# Epidemiological measures for informing the general public during the SARS-CoV-2-outbreak: simulation study about bias by incomplete case-detection

**DOI:** 10.1101/2020.09.23.20200089

**Authors:** Ralph Brinks, Helmut Küchenhoff, Jörg Timm, Tobias Kurth, Annika Hoyer

**Affiliations:** Department of Statistics, Ludwig-Maximilians-Universität Munich, Munich, Germany; Medical Faculty, Department and Hiller Research Unit for Rheumatology, University Hospital Düsseldorf, Düsseldorf, Germany; Medical Faculty, Institute of Virology, University Hospital Düsseldorf, Düsseldorf, Germany; Institute of Public Health, Charité - Universitätsmedizin Berlin, Berlin, Germany

**Keywords:** risk communication, COVID, age-structured SIR model, simulation, effective reproduction number

## Abstract

During the SARS-CoV-2 outbreak, several epidemiological measures, such as cumulative case-counts, incidence rates, effective reproduction numbers and doubling times, have been used to inform the general public and to justify interventions such as lockdown.

During the course of the epidemic, it has been very likely that not all infectious people have been identified, which lead to incomplete case-detection. Apart from asymptomatic infections, possible reasons for incomplete case-detection are availability of test kits and changes in test policies during the course of the epidemic. So far, it has not been examined how biased the reported epidemiological measures are in the presence of incomplete case detection.

In this work, we assess the four frequently used measures with respect to incomplete case-detection: 1) cumulative case-count, 2) incidence rate, 3) effective reproduction number and 4) doubling time. We apply an age-structured SIR model to simulate a SARS-CoV-2 outbreak followed by a lockdown in a hypothetical population. Different scenarios about temporal variations in case-detection are applied to the four measures during outbreak and lockdown. The biases resulting from incomplete case-detection on the four measures are compared. It turns out that the most frequently used epidemiological measure, the cumulative case count is most prone to bias in all of our settings. The effective reproduction number is the least biased measure.

With a view to future reporting about this or other epidemics, we recommend to use of the effective reproduction number for informing the general public and policy makers.

## Main Text

### Introduction

During the global SARS-CoV-2 outbreak starting in late 2019, several epidemiological measures have been used to inform the general public and policy makers. Occasionally, politicians used epidemiological measures to justify interventions such as the obligation to wear face masks or even lockdown. The most prominent epidemiological measure is the cumulative number of people tested positive, which we will call cumulative case-count (CCC). The CCC has been frequently reported on a daily basis on country level (e.g., by the COVID-19 Dashboard of the Johns Hopkins University [1]). To allow cross-country comparisons, CCC or the numbers of people tested positive often have been adjusted for population size, e.g. in the COVID-2019 situation reports of the WHO [2]. Some media reported doubling times and effective reproduction numbers. For instance, in a widely perceived press conference Germany’s Chancellor Dr. Angela Merkel declared the political goal to reach doubling times greater than ten days [3].

Whether someone is reported as an incident case, is based on the results of the diagnostic procedures carried out. Apart from anamnesis, frequently used diagnostic tests are nasopharyngeal swab tests followed by reverse transcriptase polymerase chain reaction (RT-PCR) to detect viral RNA. If the swab is done properly, RT-PCR has a moderate to high diagnostic sensitivity and a very high specificity for SARS-CoV-2 detection [4]. At least in theory, all incident cases can be identified correctly if enough screening and diagnostic efforts are spent. In practice, of course, only limited resources are available and it is likely that incident cases have been missed. Restrictions in applying diagnostic procedures are, e.g., limitations in the number of available tests kits and personnel or the local policy of running diagnostic tests. Local and even national eligibility criteria for tests have been changed during the time of the epidemic. Together with the fact that several people with the associated COVID disease show only asymptomatic or mild courses and presumably have not been considered for diagnosis of an infection at any time, the varying testing frequency leads to the hypothesis that the actual disease process was at least partially heavily underestimated. We call an infectious subject who in principle qualifies as a case but has not been detected, because she or he has not been (properly) diagnosed, an undetected case.

A useful measure for assessing case-detection is the proportion of all incident cases that were identified, i.e. the number of detected cases divided by the number of all cases (detected plus undetected cases). This proportion is referred to as the *case detection ratio* (CDR) [5].

The aim of this analysis is to determine the bias of the epidemiological measures used to inform the public due to incomplete case-detection. We conducted a simulation study using the epidemiological parameters of SARS-CoV-2 and mimic incomplete case-detection by assuming scenarios about the CDR. Next, the epidemiological measures are estimated in the presence of incomplete case-detection, and subsequently, the estimations are compared to the true values underlying the simulation.

## Methods

We use the infection-age SIR model without demography of the background host to simulate the spread of SARS-CoV-2 in a hypothetical population. In the age-structured SIR as well as in the conventional SIR model, the population is partitioned into three states, the *susceptible* state, the *infected* and the *removed* state. The initial letters of the three states give the model’s name ‘SIR’. The removed state comprises people recovered and deceased from the infected state [6]. As usual in infectious disease epidemiology, changes of numbers of people in the disease states are modeled by differential equations. Parameters of the age-structured SIR model are chosen according to the best knowledge of the SARS-CoV-2 virus. Key part of the model is the transmission rate *β*, which describes the transmission of the virus from an infectious to a susceptible individual during a given time. In the age-structured SIR model, the transmission rate depends on two time-scales: infection age *τ* and calendar time *t*. The latter is often called period in epidemiological contexts. The dependency on the infection age *τ* reflects the fact that SARS-CoV-2 transmission depends on the time since infection. For introductory texts about the SIR model, we refer the reader to [6] and the references in the electronic supplementary material.

Governments of many countries decided to invoke a lockdown, which led us to simulate three consecutive periods of the epidemic: a phase of increasing number of infections from *t* = 0 to t = 25 (days), a phase of implementation of a lockdown (from *t* = 25 to *t* = 30) followed by a phase post-lockdown when the pandemic is controlled (from *t* = 30 to *t* = 60). Start of implementation of lockdown happens at *t* = 25 (days). Figure 1 shows the marginal distributions of the transmission rate *β*. The left and right part present the dependency on the calendar time *t* and the infection-age τ.

**Figure 1:**
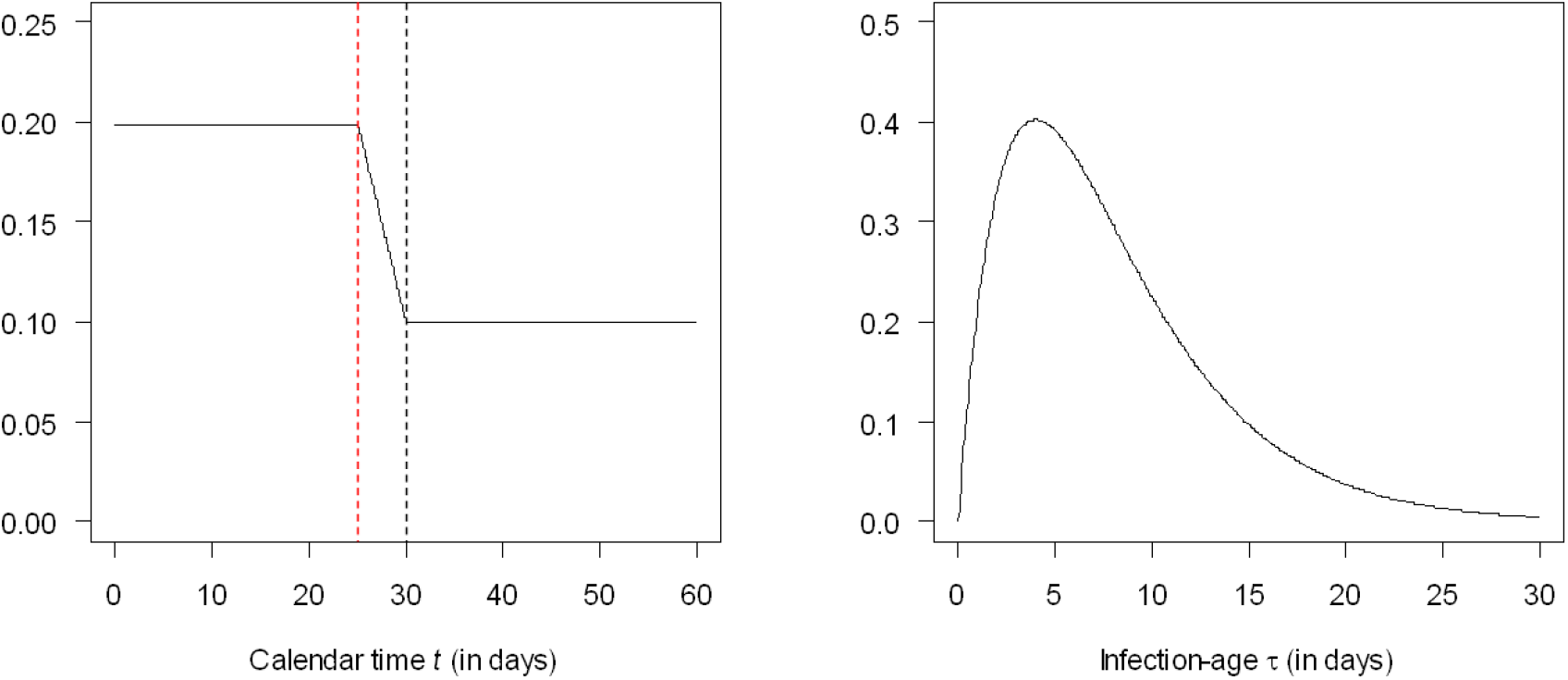
Components of the transmission rate *β*. The left panel shows the three phases of the pandemic: before lockdown at *t* = 25 (days), installation of lockdown (from *t* = 25 to *t* = 30, between vertical dotted lines) and control of the disease (*t* > 30). The right panel shows the transmission rate as a function of the infection age *τ*.

The technical details about the model as well as the source code for use in the free Statistical Software R (The R Foundation for Statistical Computing) are given in the electronic supplement.

In the case of the novel coronavirus SARS-CoV-2, new infectious cases are often reported daily [2]. If the number of newly incident cases reported on day *t* is denoted by *F*_t_ ^(o)^ (the superscipt ‘o’ indicates observed), the case-detection ratio (CDR) is the proportion of these reported cases in relation to the actual (true) but unknown number of newly incident cases *F*_t_ [5]:

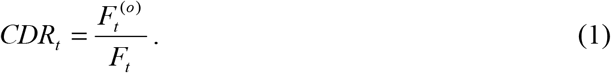

If we neglect false positive findings (as possible in highly specific tests), the CDR is a proportion ranging from 0 to 100%. In the case of a complete detection of all incident infectious cases on one day *t*, the *CDR*_t_ would equal 100% on that day.

We use different scenarios about the temporal variations in the CDR and analyze the biases imposed to epidemiological measure 1) cumulative case-counts, 2) incidence rate, 3) effective reproduction number and 4) doubling time.

1. The frequently used cumulative case count (CCC) reported on day *t* simply adds the number of incident cases until day *t*:

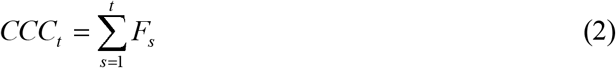 Instead of the sum of true (but unknown) incident cases, the observed *CCC*_t_^(o)^ only includes the observed number of incident cases *F*_t_^(o)^.
2. If incidence rates are reported, *F*_t_ and *F*_t_^(o)^ are usually referred to the overall size of the catchment population [2].
3. The effective reproduction number *R*_eff_ is the average number of secondary infectious cases that one primary case infects [7]. *R*_eff_ can be estimated from the number of incident cases *F*_t_. A variety of definitions of the effective reproduction number exist and we refer to the *instantaneous* reproduction number [8].
4. Sometimes, “doubling times” have been reported during the course of the pandemic. Originally, the doubling time at some point in time *t* is defined as the time span Δ = Δ(*t*) until the number of infectious cases in the population doubles [6]. As the number of infections is not easily accessible, sometimes the “doubling time” refers to the time the “cumulative incidence doubles” [9]. Thus, we use the defining condition *CCC*_t+Δ_. = 2 *CCC*_t_.

The scenarios for the temporal course of the CDR during the course of the pandemic is shown in Figure 2.

**Figure 2:**
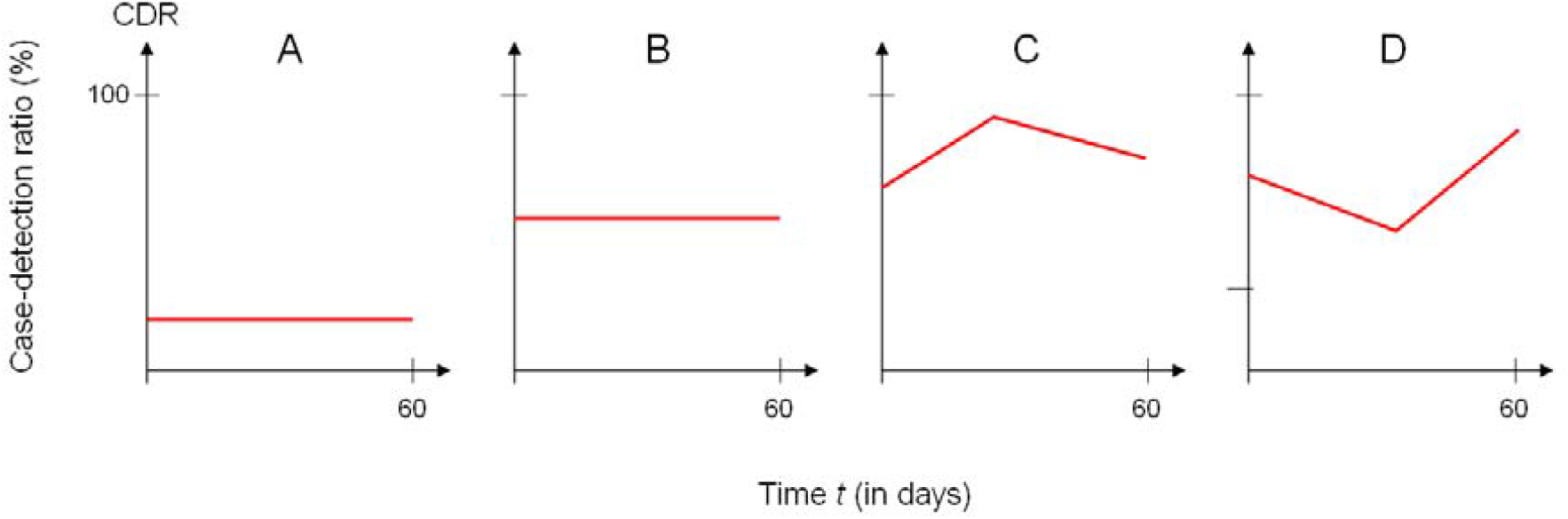
Scenarios of the time course of the case-detection ratio (CDR) during the pandemic.

In scenarios A and B we assume a low and moderate CDR without temporal trends. Scenarios C and D simulate a CDR that changes over the period of 60 days.

After running the simulation, we mimic incomplete case-detection by applying the scenarios A to D about the CDR, which will lead to incomplete case-detection. Then, the four epidemiological measures are estimated in the presence of incomplete case-detection. These estimates are compared to the true values underlying the simulation. Bias is expressed in terms of relative errors, i.e., 100% (T − E)/T, where T and E denote the true value from the simulation and the estimated value from the observed data in presence of incomplete case-detection, respectively.

## Results

Table 1 shows the biases in terms of the relative errors at days 15, 30, 45 and 60 of the pandemic. The online supplement shows the course of the relative errors during the simulated course of the epidemic (from *t* = 30 to *t* = 60).

**Table 1:** Relative error (in %) of four epidemiological measures used for informing the public during the corona pandemic.

| Measure | Scenario | Relative error (in %) |  |  |  |
| --- | --- | --- | --- | --- | --- |
|  |  | Day 15 | Day 30 | Day 45 | Day 60 |
| CCC | A | -90 | -90 | -90 | -90 |
|  | B | -50 | -50 | -50 | -50 |
|  | C | -54 | -33 | -29 | -29 |
|  | D | -18 | -27 | -27 | -27 |
| Incidence | A | -90 | -90 | -90 | -90 |
|  | B | -50 | -50 | -50 | -50 |
|  | C | -45 | -24 | -21 | -36 |
|  | D | -10 | -30 | -21 | -10 |
| Reff | A | 0 | 0 | 0 | 0 |
|  | B | 0 | 0 | 0 | 0 |
|  | C | 15 | 5 | -1 | -9 |
|  | D | -4 | -1 | 5 | 2 |
| Doubling time | A | 0 | 0 | 0 | 0 |
|  | B | 0 | 0 | 0 | 0 |
|  | C | -20 | -13 | -12 | 1 |
|  | D | 4 | 6 | -3 | -17 |

We observe that in case of the CCC and the incidence, none of the relative errors are below 10% in magnitude. In fact, relative errors of CCC and incidence can reach up to 90% in magnitude. In all examined scenarios, the effective reproduction number and the doubling time outperform the CCC and the incidence. In absolute values, the effective reproduction number is the most unbiased measure while the most frequently used CCC is the least accurate measure in the tested settings.

## Discussion

In this work we compared four epidemiological measures that have been used to inform the general public about the SARS-CoV-2 outbreak. Partly, the epidemiological measures entered the public discourse and have been used as a basis for political decisions. Changes of the measures have been used to justify interventions such as face masking, travel restrictions and lockdown. Thus, we would expect that the measures are robust with respect to the problem of incomplete case-detection. We found that particularly two measures have not proven to be robust: the frequently used cumulative case counts and the incidence rates. Instead of using these measures, for future information of the general public and policy makers, we recommend to report the relative measures “effective reproduction number” and “doubling time”.

## Data Availability

This work is based on publicly available data. All data sources have been cited properly. The source file as well as the associated technical appendix is uploaded as supplementary material.

## Declarations

### Ethics approval

The work presented here is based on a simulation study. All parameters have been cited properly in the text. No material from any human or animal have been used for this study.

### Availability of data and material

This is a simulation study. All parameters have been cited in the text.

### Code availability

The source code for this simulation study runs with the free statistical software R and is available as electronic supplementary material.

### Authors’ contributions

All authors contributed to the study conception, design, analysis and interpretation of the results. RB made the programming and wrote the first draft of the manuscript. All authors commented on previous versions of the manuscript, read and approved the final manuscript.

